# Using Networks and Prior Knowledge to Uncover novel Rare Disease Phenotypes

**DOI:** 10.1101/2025.04.02.25325098

**Authors:** Galadriel Brière, Cécile Beust, Morgane Térézol, Anaïs Baudot

## Abstract

Rare diseases are characterized by low prevalence and high phenotypic diversity. Accurately identifying phenotypes associated with rare diseases is crucial for facilitating their diagnosis and management. However, this task presents significant challenges: rare disease datasets are typically small, making statistical assessments difficult, and they often report phenotypes using various terminologies, hindering the identification of common phenotypes across datasets or the recognition of those already documented in literature and knowledge databases.

The Xcelerate RARE 2023 challenge was established to address the identification of phenotypes associated with rare diseases. Our team, MAGNET, developed a network-based approach that integrates patient clinical data from the Xcelerate RARE 2023 challenge with existing knowledge from Orphanet and the Human Phenotype Ontology (HPO). Our approach first builds a patient-disease-phenotype network comprising two layers: the Xcelerate layer encoding disease-patient-symptom associations, and the Prior Knowledge layer incorporating relationships between Orphanet rare diseases and HPO phenotypes. Then, for each rare disease included in the Xcelerate dataset, a Random Walk with Restart (RWR) algorithm is applied to the multilayer network to prioritize phenotype nodes. This framework effectively prioritizes phenotypes associated with rare diseases while distinguishing novel phenotypes from those already documented in knowledge bases, hence offering new perspectives for improving the diagnosis and characterization of rare diseases.

Our solution was awarded the prize for the *most innovative approach* in the Xcelerate RARE challenge.

## Introduction

Rare diseases often pose significant diagnostic challenges due to their low prevalence and the heterogeneity of their phenotypes (1,2). Gaining more comprehensive knowledge about phenotypes associated with rare diseases is crucial, as such information could facilitate earlier detection and more effective clinical management of these uncommon conditions.

In recent years, efforts have been made to organize the available knowledge about rare diseases. The Orphanet database (3), represents a notable example, providing a structured classification of rare diseases alongside critical information such as disease-associated genes and disease phenotypes, which are formally described using standardized terms from the Human Phenotype Ontology (HPO) (4).

The HPO plays a key role in standardizing phenotypic descriptions. However, significant challenges arise when interfacing this standardized ontology with clinical documentation practices. Patients, caregivers and clinicians often employ various terminologies that may not directly map to HPO terms. Additionally, they may describe the same symptom using HPO terms of different specificity levels within the ontological hierarchy. These inconsistencies can complicate the analysis of phenotypic patterns across rare disease cohorts, potentially limiting our understanding of rare disease associated phenotypes.

In 2023, the Xcelerate RARE challenge was established to advance the characterization of rare diseases (5,6). The initiative utilized systematically collected patient and caregiver-reported data encompassing diagnostic assessments and symptoms data, hereafter referred to as the Xcelerate dataset. The primary objective of the Xcelerate challenge was to foster the development of strategies to identify novel or underrecognized phenotype-disease associations.

We participated in the Xcelerate RARE challenge under the team name MAGNET and were awarded the prize for the most innovative solution. Our methodological framework addresses the identification of novel phenotypes associated with rare diseases through a network-based approach that explicitly incorporates the Xcelerate clinical dataset with established prior knowledge from Orphanet and HPO (hereafter referred to as the Prior Knowledge dataset). The first step of our approach consists in building a multilayer network combining disease-patient-symptom data from the Xcelerate dataset with prior knowledge on rare diseases and their associated HPO terms from Orphanet and HPO databases. Then, we formalize the detection of phenotypes associated with rare diseases as a task of node prioritization in the multilayer network. More specifically, we aim to prioritize Xcelerate symptom nodes and HPO term nodes with respect to a rare disease node of interest. To do so, we use MultiXrank (7), a Random Walk with Restart (RWR) algorithm that has shown its efficiency for node prioritization tasks on biological multilayer networks (8). The RWR algorithm explores the multilayer network starting from a selected rare disease node of interest, and assigns a score to all other nodes in the network based on how frequently they are visited during this exploration process. Hence, for a given rare disease node in the multilayer network, RWR scores reflect how strongly each phenotype node relates to the disease of interest. By ranking Xcelerate symptoms and HPO term nodes according to these scores, we can prioritize phenotypes that are most likely associated with each rare disease.

## Materials and Methods

### Xcelerate dataset

#### Disease-Patient-Symptom (XRD-Patient-Symptom) Data

The Xcelerate dataset was provided by the organizers of the challenge and is available on Synapse (6). It consists of de-identified patient data encompassing their diagnosed disease and associated patient/caregiver-reported symptoms. Overall, the Xcelerate dataset contains 741 de-identified patients affected by 27 rare diseases (Table 1). Hereafter, we will refer to the rare diseases included in this dataset as XRD (Xcelerate Rare Diseases).

**Table 1:** Number of Patients for each Xcelerate Rare Disease (XRD) in the Xcelerate Dataset and OrphaCodes of corresponding Orphanet Rare Disease (ORD), when available.

| XRD | # Patients | ORD OrphaCodes |
| --- | --- | --- |
| STXBP1 related Disorders | 79 | - |
| Wiedemann-Steiner Syndrome (WSS) | 78 | ORPHA:319182 |
| Kleefstra syndrome | 56 | ORPHA:261494 |
| FOXP1 Syndrome | 56 | ORPHA:391372 |
| CACNA1A related disorders | 52 | - |
| SYNGAP1 related disorders | 49 | ORPHA:544254 |
| CHD2 related disorders | 47 | - |
| Malan Syndrome | 45 | ORPHA:420179 |
| AHC (Alternating Hemiplegia of Childhood) | 36 | ORPHA:2131 |
| CASK-Related Disorders | 24 | - |
| DYRK1A Syndrome | 24 | ORPHA:464306 |
| HUWE1-related disorders | 23 | - |
| 8p-related disorders | 21 | ORPHA:96092 |
| CHAMP1 related disorders | 19 | - |
| 4H Leukodystrophy | 19 | ORPHA:289494 |
| Koolen-de Vries Syndrome | 16 | ORPHA:96169 |
| KDM5C-related disorders | 16 | ORPHA:85279 |
| Pallister-Killian mosaic syndrome (PKS) | 16 | ORPHA:884 |
| Classic homocystinuria | 15 | ORPHA:394 |
| Ogden Syndrome (NAA10) | 10 | ORPHA:276432 |
| MSL3 -related disorders | 9 | - |
| ARHGEF9-related disorders | 8 | - |
| Ring14 and related disorders | 7 | - |
| SETBP1-related disorder | 6 | - |
| NARS1 genetic mutation | 4 | - |
| CHOPS Syndrome | 4 | ORPHA:444077 |
| FAM177A1 Associated Disorder | 2 | - |

The Xcelerate dataset includes 222 patient/caregiver-reported symptoms across various body systems including respiratory, neurological, and growth-related symptoms (e.g. Asthma_Symptom_Present, Abnormal_Eye_Movement_Symptom_Present, Growth_Hormone_Deficiency_Symptom_Present). Most of these features are boolean (present/absent), except for 8 Children’s Sleep Habits Questionnaire (CSHQ) Subscales, for which numerical scores are reported (e.g. CSHQ Subscale 3: Sleep_Duration, CSHQ Subscale 8: Daytime_Sleepiness). We performed max normalization on the CSHQ scores (dividing each subscale by its maximum value), resulting in normalized scores between 0 and 1.

#### Xcelerate Disease-Orphanet Disease (XRD-ORD) Mappings

For 15 out of the 27 XRD contained in the Xcelerate dataset, we were provided an OrphaCode (Table 1), mapping these diseases with rare diseases included in the Orphanet database. All rare diseases included in the Orphanet database are hereafter referred to as Orphanet Rare Diseases (ORD).

### Prior Knowledge dataset

#### Disease-Phenotype (ORD-HPO) Associations

We gathered known rare disease-phenotype associations from Orphanet, available at https://www.orphadata.com/data/xml/en_product4.xml (June 2023 release). These diseases are classified within the Orphanet ontology and identified with OrphaCodes. Hereafter, we refer to these diseases Orphanet Rare Diseases (ORD).

The data we collected from Orphanet contains ORD-HPO term associations categorized by prevalence (Obligate, Very Frequent, Frequent, Occasional, Very Rare, and Excluded). We converted these prevalence categories to a uniform numerical scale from 0 to 1 (1, 4/5, 3/5, 2/5, 1/5, and 0, respectively). In total, we extracted 111,765 ORD-HPO associations, comprising 4,240 ORD and 8,305 unique HPO terms.

#### Disease-Disease (ORD-ORD) Associations

To complement the ORD-HPO associations, we computed ORD-ORD associations by leveraging Orphanet’s data on genes implicated in rare disorders (available at https://www.orphadata.com/data/xml/en_product6.xml, June 2023 release). We considered two ORDs as associated when they shared at least one common associated gene. This approach yielded 7,985 ORD-ORD associations spanning 2,771 unique ORDs.

#### Phenotype-Phenotype (HPO-HPO) Associations

We downloaded all HPO-HPO ontological associations from the HPO, available at https://hpo.jax.org/app/data/ontology (2023-06-17 release). In total, we obtained 21,556 HPO-HPO hierarchical associations.

### MultiXrank

All Random Walk with Restart computations were performed using MultiXrank (7), a python package available at https://multixrank-doc.readthedocs.io/en/latest/. We ran MultiXrank using iteratively each XRD as seed node, and using the parameter configuration described in Table 2. The global restart probability was set to 0.7. With eta=[1,0], restarts were exclusively directed to the XRD network layer, i.e. from XRD diseases and not from ORD diseases. The uniform lambda values [0.5,0.5] ensured balanced transitions between the two network layers. We configured the network layers as weighted and undirected, while interlayer connections were unweighted and undirected (graph_type=00).

**Table 2:** MultiXrank Parameters used in this study.

| MultiXrank Parameters | Value | Description |
| --- | --- | --- |
| **Network Structure** |  |  |
| Multiplex 1 | Xcelerate Layer | Contains XRD-Patient-Symptom associations. |
| Multiplex 2 | Prior Knowledge Layer | Contains ORD-HPO,ORD-ORD and HPO-HPO associations |
| Bipartite Network | XRD-ORD Interlayer Mappings | Contains XRD-ORD associations (Table 1) |
| graph_type | Multiplex 1: 01<br>Multiplex 2: 01<br>Bipartite: 00 | Network configuration codes (01: undirected weighted; 00: undirected, unweighted) |
| self_loops | 0 | Self-loops disabled |
| **RWR Parameters** |  |  |
| r | 0.7 | Global restart probability |
| delta | Multiplex 1: 0<br>Multiplex 2: 0 | Probability to change layers within a multiplex network (Set to 0 as both Multiplex 1 and 2 are monoplex networks) |
| eta | [1, 0] | Restart probability distribution across multiplex networks (in our case. restart on XRD only) |
| tau | Multiplex 1: [1]<br>Multiplex 2: [1] | Multiplex-layer-specific restart probabilities (Set to [1] as both Multiplex 1 and 2 are monoplex networks) |
| lambda | Multiplex 1 to Multiplex 2: [0.5, 0.5]<br>Multiplex 2 to Multiplex 1: [0.5, 0.5] | Inter-network transition probabilities (i.e., equal probability of staying within the Xcelerate layer or jumping to the Prior Knowledge layer, and vice versa) |

### Implementation and availability

The implementation was conducted in Python 3.10.9 as Jupyter-Notebooks to facilitate straightforward reproduction of the results. These notebooks provide comprehensive explanations for each step of the model workflow—from constructing multilayer networks using Xcelerate data and prior knowledge, to executing MultiXrank and analyzing the resulting outputs. The notebooks and results can be found on GitHub: https://github.com/BAUDOTlab/Dream_Rare-X/tree/main.

## Results

Our analysis proceeded in two main steps: first, constructing a multilayer network incorporating both patient data and established prior knowledge resources; second, applying the RWR algorithm to systematically explore and prioritize phenotype-disease associations. The following sections detail each of these steps, beginning with the construction of our Patient-Disease-Phenotype network.

### Building of the Patient-Disease-Phenotype Network

The construction of a Patient-Disease-Phenotype network, using both Xcelerate patient-derived data and Orphanet prior knowledge data sources, forms the foundation of our approach. This integration was motivated by the need to bridge the gap between clinical observations from the Xcelerate dataset and established knowledge repositories.

Our Patient-Disease-Phenotype network (Figure 1) is a multilayer network composed of two layers.

**Figure 1:**
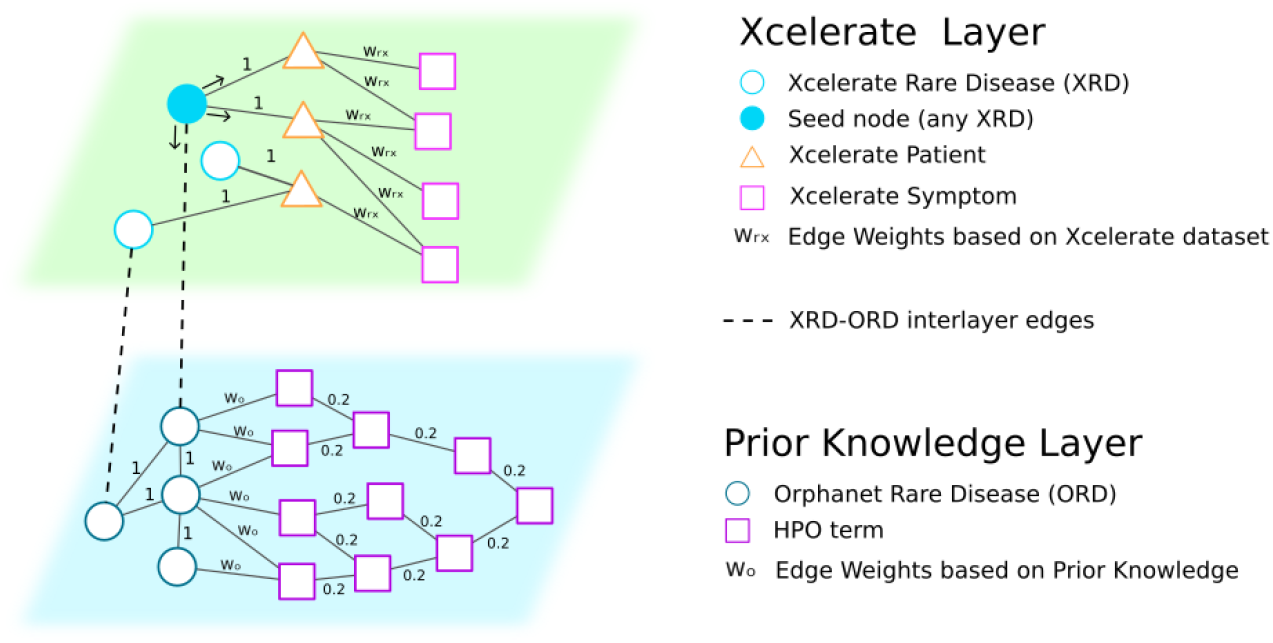
Patient-Disease-Phenotype Network

The Xcelerate layer encodes XRD-Patient-Symptom associations derived from the Xcelerate dataset (Material and Methods - Xcelerate dataset). Formally, we represented as nodes XRDs, Patients, and Symptoms. We connected Patient nodes to their diagnosed XRD using undirected edges with a weight of 1, representing a confirmed diagnostic relationship. For Patient-to-Symptom associations, we implemented two distinct weighting approaches: for boolean symptoms, we created undirected edges between Patient nodes and Symptom nodes with weight 1 when the symptom was present, establishing a binary relationship; for numerical symptoms, we created undirected edges between Patient nodes and Symptom nodes where the weight corresponded directly to the normalized value of the symptom measurement, thereby preserving the quantitative information in the network structure (Material and Methods - Xcelerate dataset, Figure 1).

The Prior Knowledge layer encodes ORD-HPO associations and ORD-ORD associations derived from public knowledge databases (Material and Methods - Prior Knowledge dataset). Formally, we represented each ORD from Orphanet and each phenotype from HPO as network nodes. We created undirected edges with weight 1 between pairs of ORD nodes when they shared at least one common associated gene, reflecting shared molecular mechanisms between diseases. We also established connections between ORD nodes and their associated HPO term nodes based on Orphanet data (Material and Methods - Prior Knowledge). Specifically, we created undirected edges between ORD nodes and HPO term nodes with weights corresponding to the prevalence of each phenotype in the respective ORD, using a scale from 0.2 to 1. Importantly, no edge was created between an ORD node and an HPO phenotype node when the phenotype was categorized as ‘Excluded’ (weight 0). Finally, we incorporated the complete phenotype ontology, connecting phenotype nodes according to the ontological structure defined in HPO. For these associations, we created undirected edges with a fixed weight set to 0.2, i.e. the minimum edge weight connecting ORD to phenotypes, corresponding to associations described as “Very Rare” in Orphanet. This specific weight choice prevents the HPO-HPO ontological relationships from dominating the information flow compared to direct ORD-HPO associations.

The Xcelerate and Prior Knowledge layers are integrated via undirected and unweighted interlayer edges, connecting 15 out of 27 XRD from the Xcelerate layer to their ORD counterpart in the Prior Knowledge layer (Table 1, Material and Methods).

### RWR on the Patient-Disease-Phenotype Network Reveals Phenotypes associated to Rare Diseases

RWR is a state-of-the-art approach for exploring networks and estimating the influence of nodes of interest (seed nodes) on other network nodes. In RWR, a random walker explores the network from a designated seed node (or a set of seed nodes), assigning scores that quantify the influence of the seed(s) on every other node. The obtained scores can be used to prioritize nodes of interest with respect to the seed(s). Recently, the MultiXrank tool has been introduced to perform RWR analysis on multilayer networks, i.e. networks composed of various layers connected to each other using interlayer interactions (7).

In this study, we used MultiXrank to perform RWR on our Patient-Disease-Phenotype network. We ran MultiXrank taking each XRD node iteratively as a seed. Starting from an XRD node, the random walker navigates within and across the two layers, overall prioritizing each node according to its relevance to the XRD seed node.

Performing RWR on our multilayer network offers three key benefits for identifying novel phenotypes and symptoms associated with rare diseases (Figure 1):

- **Leveraging clinical observations:** The random walker navigates, within the Xcelerate layer, from XRD nodes to Symptom nodes through diagnosed Patient nodes, prioritizing symptoms that are connected to the XRD by multiple paths. When many patients with a given XRD diagnosis exhibit a particular symptom, multiple paths exist between the XRD node and that Symptom node. These multiple paths increase the probability that the walker will visit and thus prioritize this Symptom node.
- **Incorporating prior knowledge:** The random walker is able to go from the Xcelerate layer to the Prior Knowledge layer using the XRD-ORD interlayer connections, allowing exploration of the Prior Knowledge layer, including the HPO hierarchy. This allows us to prioritize HPO terms directly associated with ORDs as well as their parent and broader terms in the ontology.
- **Discovering relevant HPO terms through indirect paths:** When direct XRD-ORD mappings are unavailable (Table 1), the walker can still reach the Prior Knowledge layer. Indeed, it can navigate from an XRD to Patients and Symptoms in the Xcelerate layer. Then it can move to other Patients displaying the same symptoms but diagnosed with a different XRD that has an ORD mapping. This establishes indirect paths to relevant HPO phenotypes. Additionally, by exploring ORD-ORD connections within the Prior Knowledge layer, the random walker can traverse paths between genetically related diseases. This enables the prioritization of HPO terms that may also be relevant to the XRD of interest due to common molecular mechanisms, even if not yet established in existing knowledge.

Ultimately, by comparing the top-scored Symptom nodes in the Xcelerate layer with the top-scored HPO term nodes in the Prior Knowledge layer, we expect to reveal novel symptoms associated with rare diseases that are not reported in the prior knowledge.

#### Results for XRDs with ORD mappings

For the 15 XRDs that had direct mappings to ORDs, the highest-scoring node in the Prior Knowledge layer was consistently the corresponding ORD, as expected. For these 15 XRDs, we analyzed the RWR scores as follows: we identified the 15 top-scoring Xcelerate Symptom nodes and the 15 top-scoring HPO term nodes. We manually compared these two lists to identify if the prioritized Xcelerate Symptom nodes could be associated with a corresponding prioritized HPO term. To reduce the risk of misclassifying known phenotypes as novel, for symptoms without a corresponding HPO term among the 15 top-scoring prioritized HPO terms, we extended our search to the 30 top-scoring prioritized HPO terms and to all HPO nodes linked with the corresponding ORD. Symptoms for which a corresponding HPO term could be identified were classified as consistent with known phenotypes, while symptoms without any potential corresponding HPO term were classified as potentially novel.

We illustrate as an example of manual curation the one we performed for 4H Leukodystrophy in Figure 2. The manual comparison of the two ranked lists revealed symptoms from the Xcelerate layer that directly corresponded to HPO phenotypes. For instance, the Delayed_Puberty_Symptom_Present and Short_Stature_Symptom_Present, ranked 13 and 14 respectively in the Xcelerate layer, can be matched to HPO phenotypes Delayed puberty (HP:0000823) and Short stature (HP:0004322), ranked 15 and 10, respectively, in the Prior Knowledge layer. We also estimated that the Nearsightedness_Symptom_Present (rank 7 in the Xcelerate layer) can be matched to the Myopia (HP:0000545) HPO term (rank 6 in the Prior Knowledge layer), as the two are synonyms according to HPO. We related the Coordination_Issues_Symptom_Present to the Ataxia (HP:0001251) term, as ataxia is a type of Abnormality of coordination (HP:0011443) according to the HPO hierarchy. We also matched the Cognitive_Impairment_Symptom_Present, ranked 3 in the Xcelerate later, to the Mental deterioration (HP:0001268) term, ranked 24 in the Prior Knowledge layer, as Mental deterioration is a child of the broader HPO Cognitive impairment term (HP:0100543). These HPO terms were successfully prioritized during the RWR as they are connected to the 4H leukodystrophy ODR node in the Prior Knowledge layer, indicating that the Xcelerate dataset for 4H leukodystrophy was consistent with phenotypes described in the prior knowledge.

**Figure 2:**
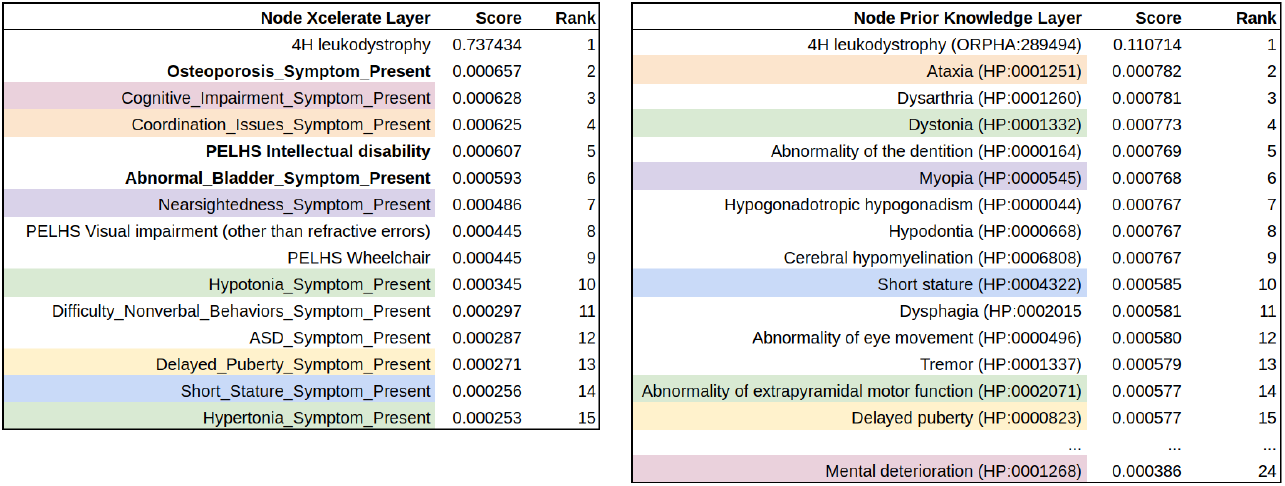
Manual curation for 4H Leukodystrophy. Colors indicate correspondences between Xcelerate symptoms and Prior Knowledge HPO phenotypes. Novel symptoms identified by our manual curation are indicated in bold.

Some other prioritized symptoms were more challenging to map to HPO terms. For instance, the symptoms Hypertonia_Symptom_Present and Hypotonia_Symptom_Present (ranked 15 and 10 in the Xcelerate layer) cannot be associated with the HPO terms, Hypotonia (HP:0001252) and Hypertonia (HP:0001276). Indeed, these HPO terms were not prioritized by the RWR because they are not considered as phenotypes associated with 4H leukodystrophy in Orphanet. However, the symptoms Hypertonia_Symptom_Present and Hypotonia_Symptom_Present can be matched with the HPO terms Dystonia (HP:0001332) and Abnormality of extrapyramidal motor function (HP:0002071). This reflects an important distinction between the clinical observations that are reported in the Xcelerate dataset and the HPO terms that are reported in Orphanet. Indeed, the Xcelerate data captures patient/caregiver-reported symptoms while Orphanet refers to etiological mechanisms. For instance, in the HPO hierarchy, Hypertonia (HP:0001276) is classified under Abnormal muscle tone (HP:0003808), which falls under Abnormality of the musculoskeletal system (HP:0033127). Meanwhile, Dystonia (HP:0001332) despite being characterized by increased muscle tone (hypertonia), is classified as an Abnormality of the nervous system (HP:0000707) rather than a musculoskeletal issue. Similarly, extrapyramidal motor system dysfunction (HP:0002071) is known to affect muscle tone regulation (9). However, this relationship between extrapyramidal motor system dysfunction and muscular tone abnormalities is not represented in the HPO hierarchy, where Abnormality of extrapyramidal motor function is classified under Abnormal central motor function (HP:0002071) within the broader Abnormality of the nervous system (HP:0000707).

Ultimately, among the top 5 symptoms prioritized in the Xelerate layer, we identified that Osteoporosis_Symptom_Present, PELHS Intellectual disability, and Abnormal_Bladder_Symptom_Present (ranked 2, 5, and 6 respectively) seem to not have corresponding prioritized HPO terms in the Prior Knowledge layer, nor were they related to any HPO term associated with the 4H leukodystrophy ORD node. Intellectual disability and Cognitive impairment are both subterms of the HPO term Abnormality of mental function (HP:0011446). While the two terms share this hierarchical relationship, they represent distinct clinical phenotypes according to HPO. Additionally, the Xcelerate dataset treats these symptoms (namely, PELHS Intellectual disability and Cognitive_Impairment_Symptom_Present) as independent features, which further justifies treating them as distinct phenotypes in our analysis. We annotated Osteoporosis, Intellectual disability and Abnormal bladder symptoms as potential novel phenotypes associated with 4H leukodystrophy. These findings are consistent with the literature on 4H leukodystrophy, as these symptoms have been previously described in several studies and case reports ((10) reports delayed bone age density and osteopenia; (11) reports osteoporosis; (12) reports osteoporosis as a long-term complication of 4H leukodystrophy; (13) and (10) report intellectual disability; (14) report intellectual deficit; (15) report chronic urinary incontinence).

Overall, the key findings from our manual curations are summarized in Table 3. The full results can be found on our GitHub repository (see Implementation and availability).

**Table 3:** Prioritized symptoms for XRD with ORD mappings. CSHQ: Sleep habit related symptoms.

| **Xcelerate Rare Disease (XRD)** | **Xcelerate symptoms consistent with known phenotypes** | **Potentially novel Xcelerate symptoms** |
| --- | --- | --- |
| 4H Leukodystrophy | Coordination Issues; Cognitive Impairment; Nearsightedness | Osteoporosis; Intellectual disability; Abnormal Bladder |
| 8P Related Disorders | Abnormal muscle function; Hypotonia | CSHQ; Heart defect |
| Alternating Hemiplegia of Childhood (AHC) | Coordination Issues; Constipation; Abnormal Eye Movement | Flexible Joints; CSHQ; Hypotonia |
| CHOPS Syndrome | Undergrowth; Short Stature; Growth Hormone Deficiency; Obesity; Asthma; Respiratory Abnormality; Decreased Pulmonary Function; COPD; Cognitive Impairment | Abnormal Muscle Function; Delivery Complication; Decreased Fetal Movement |
| Classic Homocystinuria | Heart Defect; Abnormal EKG; Bruxism; Feeding Difficulties | CSHQ; Abnormal Muscle Function; Cafe Au Lait Spots |
| DYRK1A Syndrome | ASD; Repetitive Stereotypic Behavior; Undergrowth; Short Stature; Abnormal Muscle Function; Esophagus Issues; Feeding Difficulties | CSHQ; Unusual Eye Gaze; Abnormal Optic Nerve |
| FOXP1 Syndrome | Cognitive Impairment; ASD; Short Attention Span; Abnormal Eye Movement; Unusual Eye Gaze; Learning disability; Coordination Issues | Impulsivity; Constipation; Hypotonia; Abnormal Muscle Function; Sleep Anxiety |
| KDM5C-related disorders | Impulsivity; Short Attention Span | CSHQ; Abnormal Bladder; Anxiety; Cognitive impairment |
| Kleefstra syndrome | Cognitive impairment; Coordination issues; Hypotonia; Abnormal eye movement; Abnormal ear symptom | Constipation; Feeding Difficulties |
| Koolen-de Vries Syndrome | Hypotonia; Cognitive impairment; Coordination issues | Abnormal EEG; Seizures; Immunodeficiency; Autoimmune disorder |
| Malan Syndrome | General overgrowth; Cognitive impairment; Muscles and bones issues; Eye issues | Coordination issues; CSHQ |
| Ogden Syndrome (NAA10) | Abnormal muscle; Hypotonia; Issue muscle arms legs; CSHQ Subscale 8: Daytime Sleepiness; PELHS Intellectual disability | Cognitive impairment, Coordination issues; Hyperactivity; Short attention span |
| Pallister-Killian mosaic syndrome (PKS) | Cognitive impairment; Hypotonia; Pigmentation issues; Coordination issues; Abnormal Inside Mouth; Abnormal Hair | Constipation; Abnormal EEG; Seizures |
| SYNGAP1 related disorders | ASD; Abnormal muscle function; Cognitive impairment; Hypotonia; Temper tantrums; Aggressive Violent Behavior; Impulsivity | Eye gaze; Abnormal EEG; Intellectual disability; Seizures |
| Wiedemann-Steiner Syndrome (WSS) | Hypotonia; Abnormal muscle function; Temper tantrums; Aggressive violent behavior; Anxiety; Feeding difficulties; Constipation | Short stature; Undergrowth; Cognitive impairment |

#### Results for XRD without ORD mappings

For the 12 XRD lacking a direct mapping to ORD (Table 1), our analysis revealed that Wiedemann-Steiner syndrome ORD always received the highest scores in the Prior Knowledge layer. We attribute this high scoring to three key factors:

- **Reachability from the Xcelerate layer:** The Wiedemann-Steiner syndrome XRD has a corresponding ORD in the Prior Knowledge layer, so the ORD node for the syndrome can easily be reached through the interactions connecting the two layers.
- **High representation in the Xcelerate dataset:** The Wiedemann-Steiner syndrome is largely represented in the Xcelerate cohort, with a total of 78 patients diagnosed with this condition (second most represented disease in the cohort, Table 1).
- **Numerous unspecific symptoms:** This syndrome exhibits numerous unspecific symptoms within the dataset. Indeed, the symptoms associated with Wiedemann-Steiner syndrome XRD are also found in many other XRD. For example, the “Short_Stature” symptom is associated with 21 out of the 27 diseases in the Xcelerate layer; the “Undergrowth_Symptom_Present” is linked to 20 diseases, and “Hypotonia_Symptom_Present” is linked to 26 diseases. Therefore, the Wiedemann-Steiner syndrome XRD node in the Xcelerate layer can easily be reached through any of these unspecific symptoms. Once the syndrome has been reached in the Xcelerate layer, it can diffuse to the ORD node via the interlayer interaction.

In summary, due to these 3 factors, the Prior Knowledge scores obtained for the 12 XRD without mapping should be interpreted with caution. However, the scores of nodes from the Xcelerate layer do not suffer from this limitation. Hence, it is still possible to interpret high-scoring Symptom nodes from the Xcelerate layer.

## Discussion

Our study demonstrates the effectiveness of network-based approaches for elucidating phenotype-disease associations in rare diseases. The RWR algorithm applied to our Patient-Disease-Phenotype network has successfully identified both established and potentially novel phenotypes associated with rare diseases in the Xcelerate dataset. This approach addresses several critical challenges in rare disease characterization.

First, the integration of patient/caregiver-reported data from the Xcelerate dataset with established knowledge repositories (Orphanet and HPO) creates a comprehensive framework that bridges the gap between clinical observations and standardized ontologies. This integration is particularly valuable in the context of rare diseases, where limited patient cohorts and terminological inconsistencies can hinder systematic phenotypic characterization. Moreover, the incorporation of disease-disease relationships based on shared genetic etiology in the Prior Knowledge layer facilitates knowledge transfer between genetically related conditions.

Second, our model is able to prioritize symptoms with high specificity, even when they’re rare in the target disease cohort. This behaviour is exemplified in the case of 4H leukodystrophy, where our analysis yielded a seemingly paradoxical result: osteoporosis, with minimal prevalence in the 4H leukodystrophy cohort (present in only 2 patients out of the 19 diagnosed with 4H leukodystrophy) and across the full cohort (present in only 7 patients total out of the 741 included in the cohort), received a higher prioritization score than cognitive impairment or coordination issues (present in 6 patients each in the 4H leukodystrophy cohort and in 302 and 273 patients respectively in the full cohort). This prioritization of osteoporosis over cognitive impairment and coordination issues can be explained through examination of the RWR parameters and their interaction with the symptom-disease network topology. The high restart probability (r=0.7) used for the RWR influences the network exploration. For specific symptoms such as osteoporosis, the random walker is less likely to explore other diseases’ neighborhoods, as few paths in the network connect the osteoporosis symptom to patients diagnosed with other diseases. The combination of this topology with the high restart probability makes the random walker more likely to stay within close range of the osteoporosis node, once it’s reached, before a restart event. Conversely, symptoms that are shared across many diseases, such as cognitive impairment and coordination issues, establish multiple connections to diverse disease nodes. The walker is more likely to explore other diseases’ neighborhoods before a restart event, thus diluting the scores of these common Symptom nodes. This behavior allows the RWR to highlight symptoms with discriminative value between disorders, even when their frequency within the target cohort is low. By prioritizing symptoms with higher diagnostic specificity, our approach could provide a valuable tool uncovering distinctive clinical signatures in rare disorders.

However, our study presents several limitations. First, while we obtained good results for Xcelerate diseases that had a disease mapping in Orphanet, we observed that the results in the Prior Knowledge layer should be interpreted with caution for the diseases lacking an ORD mapping. However, for those diseases, the results from the Xcelerate layer can still be exploited to prioritize Symptom nodes from the Xcelerate layer. Second, the interpretation of the results is currently done manually. An automation of this process would be valuable. To go in this direction, we propose to include interlayer relations associating Xcelerate symptoms to corresponding HPO terms. This would both benefit the RWR scores, allowing for a direct information flow from HPO nodes to Xcelerate symptoms nodes, and the automatic comparison of symptoms and phenotypes prioritized by the model. However, this mapping between symptoms and associated HPO is far from trivial. As demonstrated in our analysis of 4H leukodystrophy, there exists a fundamental distinction between symptoms reported in patient/caregiver-reported clinical studies and the phenotypic descriptions found in reference resources such as Orphanet, which tend to emphasize etiological mechanisms. Additionally, while HPO contains both symptomatic and etiological terms, it lacks explicit causal relationships between them (e.g., the relation between extrapyramidal dysfunction and abnormalities of muscle tone). This challenge makes automatic mapping between patient-derived symptoms and HPO term particularly difficult. Incorporating causal relationships directly in the Prior Knowledge layer would significantly enhance the model’s ability to connect phenotypes with their underlying mechanisms. These limitations underscore the necessity for expert manual curation in the current workflow, while also highlighting the need for developing automated systems capable of bridging the gap between patient/caregiver-reported symptoms and their representation in standardized knowledge resources.

## Conclusion

In this study, we presented a network-based approach that integrates patient/caregiver-reported data with established knowledge repositories to improve the phenotypic characterization of rare diseases. Our method, which earned the most innovative solution award in the Xcelerate RARE challenge, successfully identified both known and potentially novel phenotype-disease associations through a Random Walk with Restart algorithm on an multilayer patient-disease-phenotype network. This work demonstrates the value of network approaches for rare disease characterization and offers a promising methodological framework to support discoveries in rare disease research. Our solution is available as well-documented Jupyter notebooks, enabling easy reuse for users who wish to reproduce our results or apply the methodology to their own data, from network construction to RWR execution and result interpretation.

## Data Availability

All data used in this study have been obtained from the Xcelerate RARE Open Science Data Challenge. The data can be accessed with a registered account on Synapse:
SB. RARE-X A Rare Disease Open Science Data Challenge [Internet]. [cited 2025 Mar 17]. Available from: https://www.synapse.org/Synapse:syn51198355

https://www.synapse.org/Synapse:syn51198355

## Acknowledgments

This work was supported by the French National Research Agency (ANR-21-CE45-0001), the European Rare Diseases Research Alliance (ERDERA), the MarMaRa Institute (AMX-19-IET-007), and the European Union’s Horizon 2020 research and innovation programme (EJP RD COFUND-EJP No 825575).

